# Assessing Real-Life Food Consumption in Hospital with an Automatic Image Recognition Device: a pilot study

**DOI:** 10.1101/2024.10.04.24314889

**Authors:** Laura Albaladejo, Joris Giai, Cyril Deronne, Romain Baude, Jean-Luc Bosson, Cécile Bétry

**Author notes:** Corresponding author: Laura Albaladejo.

## Abstract

**Background and aims:** Accurate dietary intake assessment is essential for nutritional care in hospitals, yet it is time-consuming for caregivers and therefore not routinely performed. Recent advancements in artificial intelligence (AI) offer promising opportunities to streamline this process. This study aimed to evaluate the feasibility of using an AI-based image recognition prototype, developed through machine learning algorithms, to automate dietary intake assessment within the hospital catering context.

**Methods:** Data were collected from inpatient meals in a hospital ward. The study was divided in two phases: the first one focused on data annotation and algorithm’s development, while the second one was dedicated to algorithm’s improvement and testing. Six different dishes were analyzed with their components grouped into three categories: starches, animal protein sources, and vegetables. Manual weighing (MAN) was used as the reference method, while the AI-based prototype (PRO) automatically estimated component weights. Lin’s concordance correlation coefficients (CCC) were calculated to assess agreement between PRO and MAN. Linear regression models were applied to estimate measurement differences between PRO and MAN for each category and their associated 95% confidence intervals.

**Results:** A total of 246 components were used for data annotation and 368 for testing. CCC values between PRO and MAN were: animal protein sources (n= 114; CCC = 0.845, 95% CI: 0.787-0.888), starches (n= 219; CCC = 0.957, 95% CI: 0.945-0.965), and vegetables (n=35; CCC = 0.767, 95% CI: 0.604-0.868). Mean differences between PRO and MAN measurements were estimated at -12.01g (CI 95% -15.3, -8,7) for starches (reference category), 1.19 g (CI 95% -3.2, 5.6) for animal protein sources, and -14.85 (CI 95% -22.1, -7.58) for vegetables.

**Conclusion:** This pilot study demonstrates the feasibility of utilizing an AI-based system to accurately assess food types and portions in a hospital setting, offering potential for routine use in clinical nutrition practices.

## Introduction

Assessing dietary intake is essential for delivering optimal nutritional care to inpatients (1–3). This process facilitates the identification of malnutrition risks, the monitoring of nutritional intake, and the customization of dietary interventions based on individual patient needs. Standard methods for dietary assessment include food diaries, 24-hour dietary recalls, and food frequency questionnaires (FFQs) (1,4). However, these methods are time-consuming and rely on patients’ memory and cooperation, which can be challenging for individuals with cognitive impairments, fatigue, or acute illnesses (5). Furthermore, such approaches are susceptible to various biases, such as recall bias, social desirability bias, and measurement errors, which can undermine the accuracy and reliability of the data collected (6). Alternatives, such as direct observation during meals-where caregivers visually estimate food consumption- or the pre- and post-meal weighing of food to determine actual intake, are possible but rarely implemented due to constraints on time and personnel availability.

Innovative methods using artificial intelligence (AI) have been developed to automate the assessment of food intake (7,8). These methods involve food segmentation, classification, and volume estimation. However, this process is inherently challenging because it requires multiple images, controlled environments, and specialized equipment, making it highly sensitive to image acquisition conditions, which can ultimately affect accuracy (9). These approaches have predominantly been tested in laboratory settings, which may limit their robustness and applicability in real-world environments, particularly in hospitals (8). The challenge of accurately assessing food intake in care facilities remains underexplored. One approach has been developed to automatically recognize food on hospital meal trays and estimate its quantity. This technology demonstrates efficacy in food recognition, but has not been validated for accurate food quantification, likely due to its reliance on two-dimensional images (10). Another study in long-term care homes highlighted difficulties in accurately estimating food volume (11). Furthermore, although a study in older inpatients demonstrated good reliability, it relied on visual estimation as the ground truth, which introduces inherent limitations and potential inaccuracies (12). The aim of our study was to train and assess a prototype based on machine learning algorithms for assessing real-life food consumption in hospitals.

## Materials and methods

### Data collection

Measurements were conducted on the main courses of the standard hospital diet over three consecutive days, covering both lunch and dinner in a hospital ward. For each meal tray, only the main course was measured, with trays either being test trays measured before consumption or trays collected post-meal. Meals in our facility are served in sealed white plastic trays, with the seals removed before measurements. Data for each of the six dishes were taken three times, once every three weeks over three cycles, in line with the hospital’s three-week menu rotation. This resulted in three distinct datasets and ensured consistency in the dishes analyzed (Figure 1). The first dataset was used for data annotation and algorithm development (phase 1), while the second and third datasets were used for algorithm improvement and testing (phase 2). Depending on the meal, dishes could contain either a single component (e.g., pizza) or multiple components (e.g., minced beef steak with pilaf rice and piperade). Six different dishes were analyzed: royal pizza, ricotta and spinach tortellini, potato tortilla, hake with piperade and pilaf rice, minced beef steak with pasta, and smoked sausage with wheat berries and vegetable brunoise (Figure 1). Each component was classified according to its primary nutrient contribution. For example, although pizza contains some ham, it was categorized as a starch. This classification resulted in three categories: animal protein sources (hake, minced beef steak, and smoked sausage), starches (pilaf rice, pasta, wheat berries, potato tortilla, ricotta and spinach tortellini, and royal pizza), and vegetables (piperade and brunoise).

**Figure 1.**
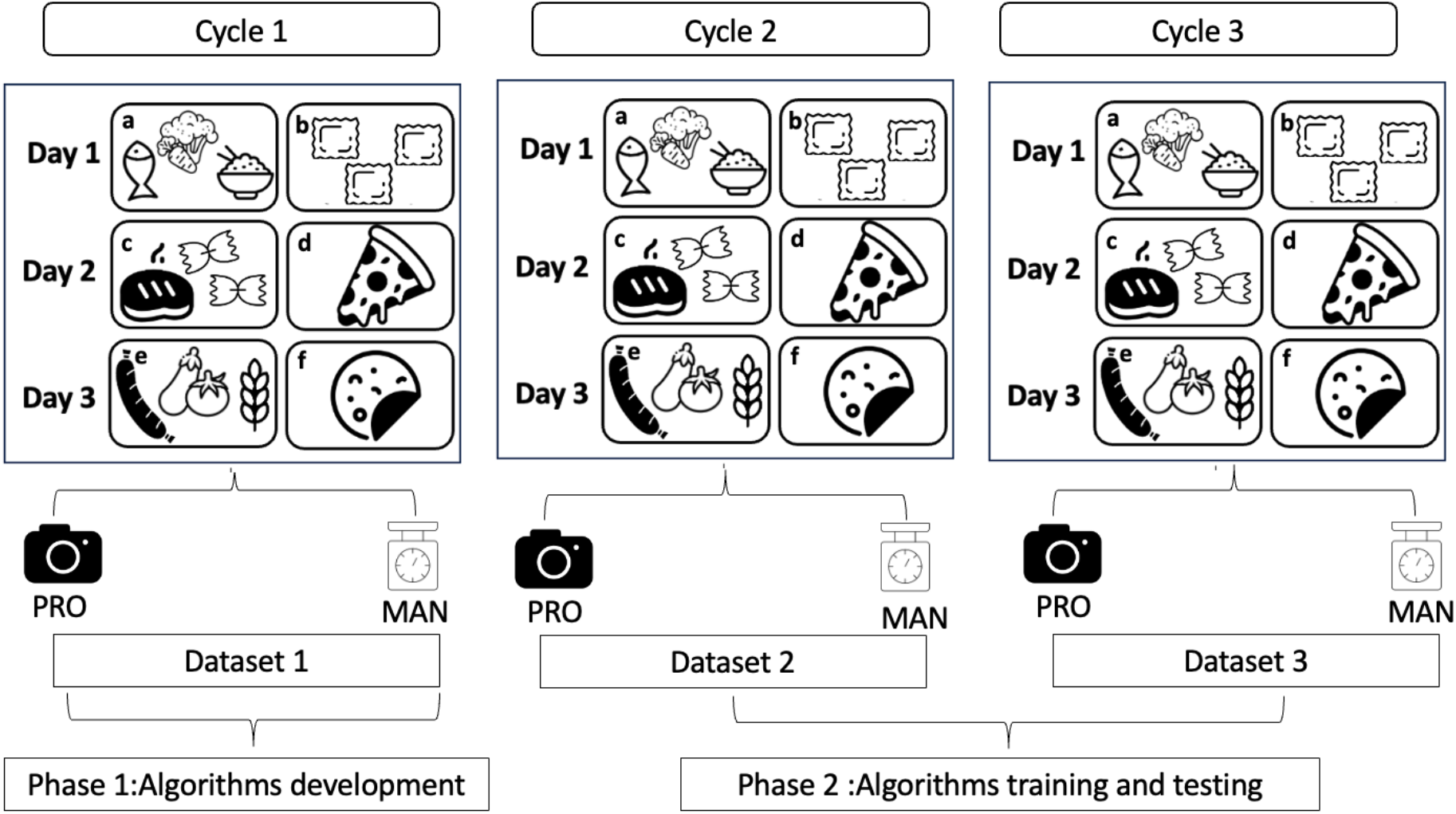
Data collection methodology *a: hake with piperade and pilaf rice* *b* : *ricotta and spinach tortellini* *c: minced beef steak with pasta* *d* : *royal pizza* *e: smoked sausage with wheat berries and vegetable brunoise* *f: potato tortilla*

#### The manual method (MAN)

For manual weighing, each component of the main courses was weighed separately by a registered dietitian (LA) using the same electronic scale (GPISEN Smart Digital Balance, precision ± 1 g, China). Weighing was performed after collection of data with the prototype.

#### The automatic image recognition device prototype (PRO)

The prototype was designed by DMCC and APREX Solutions (Nancy, France). It consisted of 1) an image acquisition unit and 2) an image processing unit, which also displayed and recorded data. The prototype was designed to collect data in approximately one second, without any operator interaction. The system was automatically triggered when a tray was presented to the device (Figure 2A and 2B). The image acquisition unit consisted of a 2D camera and a stereoscopic camera, both of which provided accurate 3D reconstructions of the food’s color map (Figure 2C). The cameras were controlled by AX Vision software (APREX Solutions, Nancy, France) which managed image capture, exposure settings, field of view, and the 3D parameters of the stereoscopic camera. Once the 3D and 2D images were captured, a set of algorithms was applied to estimate the volume of the food. These algorithms were based on an improved DenseNet-type neural network (AX IA Software, APREX Solutions, Nancy, France) The weight estimation was derived from measurements obtained from the first dataset.

**Figure 2.**
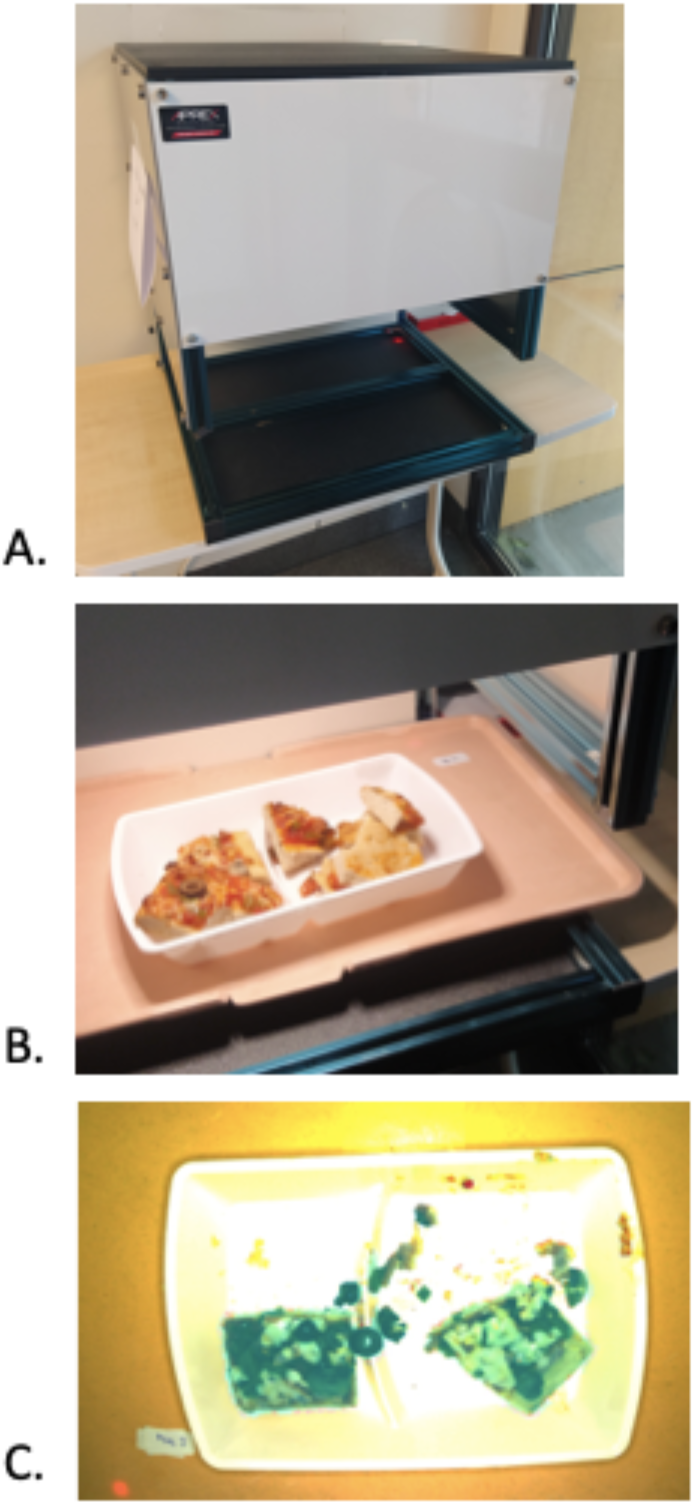
The prototype (A), meal tray positioning (B), and image capture process (C)

### Statistical analyses

Quantitative data were presented as means and standard deviations after verifying the distribution graphically. Lin’s Concordance Correlation Coefficients (CCC) were calculated for each category-animal protein sources, starches, and vegetables -to evaluate the agreement between PRO and MAN. This analysis assessed how closely the measured values aligned with the line y = x, which represents perfect agreement between the two measurements methods considering MAN was the gold standard. The interpretation of CCC followed Partik (2002). Linear regression models were fitted on the difference between PRO and MAN measurements ((MAN-PRO), dependent variable) separately for each category (predictor). Corresponding marginal means were then estimated and displayed with their 95% confidence intervals. All tests were two-tailed and considered significant at the 5% level. Statistics were performed using Jamovi software (version 2.3) (15).

## Results

### Data collection

We collected in the algorithm development phase, 246 components data from 92 images (dataset 1) and in the testing phase 368 components data from 183 images (datasets 2 and 3). The statistical analysis is based on datasets 2 and 3.

### Concordance correlation coefficients

Lin’s concordance correlation coefficients between the two measurement tools were as follows for each component: animal protein sources (CCC = 0.845, 95% CI: 0.787-0.888), starches (CCC = 0.957, 95% CI: 0.945-0.965), and vegetables (CCC = 0.767, 95% CI: 0.604-0.868) corresponding to fairly good, excellent and moderately satisfactory concordance respectively. The CCC for animal protein sources indicated fairly good concordance (0.81-0.90), while for starches, the coefficient suggested excellent concordance (>0.95). In contrast, the CCC for vegetables was categorized as moderately satisfactory (0.71-0.80). (Figure 3).

**Figure 3.**
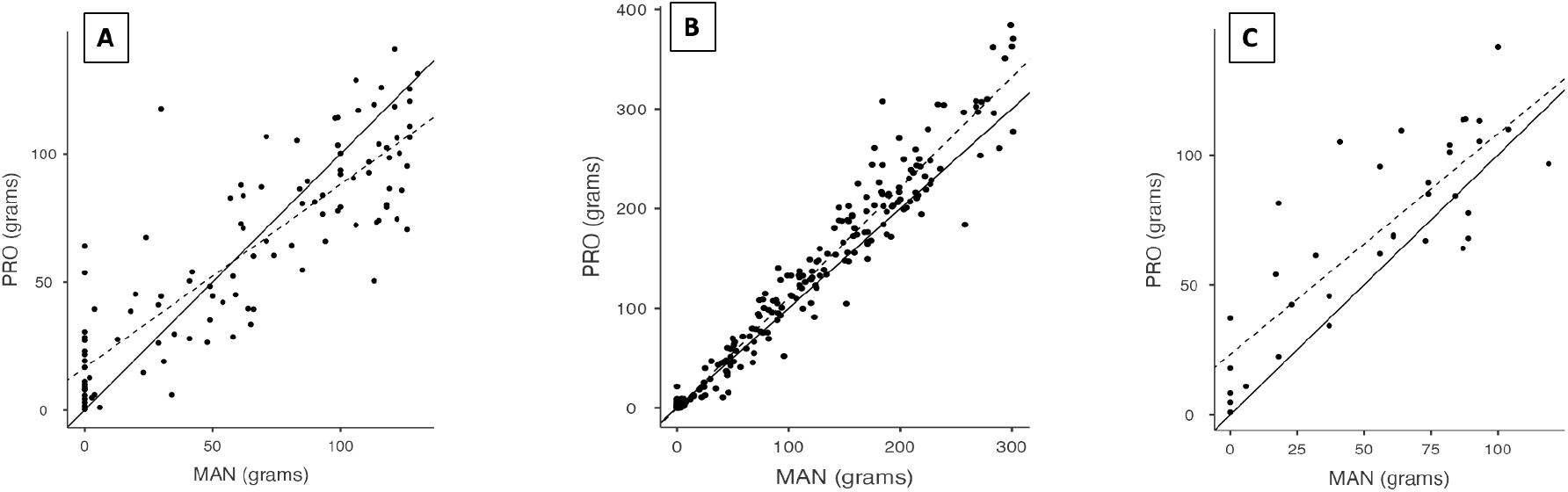
Concordance correlation coefficient between the automatic image recognition device prototype (PRO) and the manual weighing method (MAN) for animal protein sources (**A**, n=114), starches (**B**, n=219) and vegetables (**C**, n=35).

The solid line represents the *x = y* line, indicating perfect agreement. The dotted line represents the linear regression between PRO and MAN. A CCC of 1 indicates perfect overlap of the two lines.

### Linear Regression Models

The estimated mean differences between PRO and MAN measurements are as follows: -12.01 g (CI 95% -15.2, -8.8) for starches (reference category), 1.19 g (CI 95% -3.2, 5.6) (calculated as - 12.01 + 13.2) for animal protein sources and -14.85 g (CI 95% -22.8, -6.9) (calculated as -12.01 - 2.84) for vegetables. PRO overestimates the weight compared to MAN for both starches and vegetables (P < 0.001) (Table 1).

**Table 1.** Difference (linear regression model) in measurement estimates between the automatic image recognition device prototype (PRO) and the manual weighing method (MAN) by food type, n= 368.

| Predictor | Estimate | SE(Estimate) | p |
| --- | --- | --- | --- |
| Intercept | -12.01 | 1.61 | <0.001 |
| **Type of food (reference: Starches):** |  |  |  |
| Animal protein sources | 13.20 | 2.77 | <0.001 |
| Vegetables | -2.84 | 4.35 | 0.514 |

## Discussion

The primary aim of this study was to evaluate the accuracy of an automatic image recognition device prototype that utilizes optical and artificial intelligence (AI) algorithms for estimating food quantities on a meal tray. The findings demonstrate that the prototype is reasonably accurate in estimating food weights, particularly for animal protein sources and starches. Lin’s concordance correlation coefficients indicate a high level of agreement between the prototype and manual measurements for these categories, suggesting that the device could potentially replace manual methods under certain conditions. However, for vegetable portions, the prototype shows less reliable accuracy, with higher average errors of 15 grams compared to 1 gram for animal protein sources and 12 grams for starches. This discrepancy may be attributed to the varied shapes and textures of vegetables, which present greater challenges for image recognition and volume estimation.

There has been an increasing number of AI-based tools using imaging for the automated analysis of nutritional intake. Many of these tools, such as the “Foodintech” solution developed for inpatients, rely on two-dimensional (2D) images captured via smartphones mounted on meal carts (10). While these tools have shown some success in recognizing dishes, their capability to quantify food intake remains limited, mainly due to the inability of 2D images to capture food thickness and volume accurately. Public mobile applications face similar limitations, where the nutritional assessments can vary widely compared to traditional 3-day food diary (16). In contrast, systems utilizing three-dimensional (3D) models offer more precise food intake assessments by accounting for the volume and density of food items (17–20). Our prototype’s performance aligns with these findings, showing a mean percentage error for portion size estimation that is consistent with values reported in the literature (21)(22)(23) (24).

We acknowledge limitations in our research. First, the manual weighing was conducted by a single dietitian, which may introduce operator bias. Future studies should include multiple operators to reduce this bias and enhance data robustness. However, a key strength of this study is the direct comparison of the prototype with a calibrated kitchen scale, providing a more reliable reference than the visual estimation commonly used in clinical settings. A second limitation involves the detection errors related to sauces, which the current algorithm incorrectly categorizes as animal protein sources. Refining the algorithm to better identify and classify sauces could improve overall accuracy. Lastly, the prototype’s performance was affected by the hospital meal trays, particularly when food was stuck in tray grooves. Addressing these packaging-related challenges, perhaps through design modification or alternative imaging techniques, is essential for more accurate assessments.

Several perspectives are envisioned for further development of the prototype. Indeed, several sources of error in energy intake estimation could occur, notably due to the use of volume-based data from a database for mass estimation, which is then used for caloric calculations. One prospect is to integrate macronutrient data to enable complete analyses of calorie, protein, lipid, and carbohydrate intake. This can be achieved by incorporating food composition tables and recipes supplied by the catering industry into the technology (25). Finally, the prototype is currently heavy and difficult to transport. Improving its ergonomics and co-constructing it with caregivers will be necessary to ensure it meets their needs and work organization.

## Conclusion

Food intake assessment is essential for the prevention and management of malnutrition in hospitals. We developed an automatic image recognition device prototype that is reliable for automatic food volume and weight recognition. This new technology represents a valuable tool to help quantify food intake in healthcare departments. In the long term, using such a tool could enable accurate quantification of hospitalized patients’ food intake and save paramedical staff time.

## Data Availability

All data produced in the present study are available upon reasonable request to the authors

## Funding statement

DMCC company financed the developpement of the prototype. The project was financially supported by the Société Francophone de Nutrition Clinique et Métabolisme (SFNCM).

## Conflict of interest

LA, JG, JLB and CB declare no conflict of interest

CD is manager of the company that designed the prototype. RB is the manager of the company that designed the prototype’s algorithms.

## Author contributions

CD and RB conceived the prototype. LA, RB, JLB and CB designed the study. LA collected the data. LA, JG, RB and JLB analyzed the data. LA, JG, JLB and CB contributed to the interpretation of the results. LA and CB wrote the first draft. All authors discussed the results and contributed to the final manuscript.

## Notes

### Funding Statement

DMCC company financed the developpement of the prototype. The project was financially supported by the Societe Francophone de Nutrition Clinique et Metabolisme (SFNCM).

